# Expectant parents’ perceptions of healthcare and support during COVID-19 in the UK: A thematic analysis

**DOI:** 10.1101/2021.04.14.21255490

**Authors:** Ezra Aydin, Kevin A. Glasgow, Staci M. Weiss, Topun Austin, Mark Johnson, Jane Barlow, Sarah Lloyd-Fox

## Abstract

**Background:** In response to the COVID-19 pandemic, expectant parents experienced changes in the availability and uptake of both NHS community and hospital-based healthcare.

**Objective:** To examine how COVID-19 and its societal related restrictions have impacted the provision of healthcare support for pregnant women during the COVID-19 pandemic.

**Method:** A thematic analysis using an inductive approach was undertaken of data from open-ended responses using data from the national COVID in Context of Pregnancy, Infancy and Parenting (CoCoPIP) Study online survey (N = 507 families).

**Results:** The overarching theme identified was the way in which the changes to healthcare provision increased parents’ anxiety levels, and feelings of not being supported. Five sub-themes, associated with the first wave of the pandemic, were identified: (1) rushed and/or fewer antenatal appointments, (2) lack of sympathy from healthcare workers, (3) lack of face-to-face appointments, (4) requirement to attend appointments without a partner, and (5) requirement to use PPE. A sentiment analysis, that used quantitative techniques, revealed participant responses to be predominantly negative (50.1%), with a smaller proportion of positive (21.8%) and neutral (28.1%) responses found.

**Conclusion:** This study provides evidence indicating that the changes to healthcare services for pregnant women during the pandemic increased feelings of anxiety and have left women feeling inadequately supported. Our findings highlight the need for compensatory social and emotional support for new and expectant parents while COVID-19 related restrictions continue to impact on family life and society.

## Introduction

In March 2020 the World Health Organisation (WHO) declared a global pandemic stemming from the emergence of a novel coronavirus, SARS-CoV-2, resulting in the potentially fatal disease COVID-19. This led to the rapid implementation of new policies by many governments to contain and mitigate the spread of infection. In response to the social distancing requirements put in place by the UK Government to curtail the spread of infection, Public Health England (PHE) as well as similar bodies in the devolved nations (Scotland, Wales and Northern Ireland) instigated guidance regarding the delivery of community and hospital-based NHS services that involved their cessation or delivery virtually (i.e., telephone or video call). Antenatal and maternity services available to expectant parents have been consistently reported as being disproportionately affected by these recommendations (Human Rights in Childbirth, 2020). Regional variations in healthcare provision and advice for expectant mothers have also been reported, including birth partners being denied access to the hospital, and limited access to babies admitted to neonatal intensive care (Karavadra et al., 2020). Most worrying is the suggestion of a increase in stillbirths observed in a sample of 1681 births in London, UK between February and June 2020 (Khalil et al., 2020). Authors attributed this increase to lack of preventive antenatal care (Khalil et al., 2020).

A number of studies examining women’s subjective experiences of pregnancy-based services confirmed these reports. For example, in May 2020, a study observing the healthcare experiences of expectant women in the UK found that 59% of respondents to the online survey perceived barriers to accessing healthcare at the time of their pregnancy during the initial COVID-19 lockdown (Karavadra et al., 2020). These included changes in the way services are delivered (i.e., virtual), lack of information provided during routine appointments, and reluctance to discuss mental health issues virtually. Similarly, an online study of 5,474 families between 23rd March - 4th July 2020, found that 38% of participants were concerned about their ability to get reliable pregnancy information and advice during this time (Best Beginnings, Home-Start UK, and the Parent-Infant Foundation UK, 2020).

Heightened anxiety and depression have also been reported during the initial national lockdown (Pierce et al., 2020), with expectant and new mothers (with an infant under the age of one) experiencing unique physical and psychological stressors (Davenport et al., 2020). Recent studies found stress in expectant parents was partially attributed to changes in the access and medium (virtual, on the phone, etc.) of medical support and perinatal services (Karavadra et al., 2020; Reingold et al., 2020). Additional stressors related to the wider secondary social and economic impact of the pandemic on family life (i.e., in terms of financial problems, limited support from families and wider services, and the additional burdens of home-schooling etc., Ahlers-Schmidt et al., 2020; Chivers et al., 2020), in addition to the impact of media messages concerning the safety of seeking medical or midwifery help during the pandemic (e.g. fear of contracting COVID-19 on route to, or whilst inside, a hospital attending a routine appointment (Fakari & Simbar, 2020)).

The COVID-19 in the Context of Pregnancy, Infant Parenting (CoCoPIP) Study was developed to explore how COVID-19 and the cascade of changes in healthcare, social restrictions and government guidance impacted the lives of families who were expecting a baby or had recently given birth. The aim of the current analysis is to explore expectant family’s perceptions of their healthcare appointments, health and social support (i.e., access for partner to attend visits) during COVID-19, and to provide insight into potential barriers women have experienced in their antenatal care, giving a voice to expectant parents in the UK.

## Methods

### Participants

Survey data is taken from the period 14th July - 5th December 2020 (n = 507, see *Table 1* for demographic information). Recruitment strategies included contacting nationwide antenatal and postnatal health groups directly, social media platforms (Twitter, Facebook and Instagram), as well as other child development research groups and networks in the UK. Eligibility criteria for the study were expectant parents (who had completed their first trimester) or parents of an infant between the ages of 0-4 months, who were then asked to report on experiences during their recent pregnancy. All participating parents gave informed consent to take part in the CoCoPIP online survey (<u>tinyurl.com/CoCoPIP</u>). Ethics approval for the survey was given by the University of Cambridge, Psychology Research Ethics Committee (PREC) (PRE.2020.077).

**Table 1:** Participant demographic information

| Demographic | Proportion n |
| --- | --- |
| **Who** |  |
| Mother | 503 |
| Father | 2 |
| Non-birth Mother | 1 |
| Other partner | 1 |
| **Ethnicity** |  |
| White | 464 |
| Black | 9 |
| Asian | 10 |
| Mixed/multiple | 10 |
| Hispanic | 9 |
| Arab | 0 |
| Other | 2 |
| Undisclosed | 12 |
| **Index of multiple deprivation (IMD)** |  |
| MD (most deprived) | 9 |
| 2 | 21 |
| 3 | 25 |
| 4 | 32 |
| 5 | 25 |
| 6 | 43 |
| 7 | 32 |
| 8 | 45 |
| 9 | 38 |
| LD (least deprived) | 34 |
| Postcode not identified | 35 |
| Undisclosed or partial postcode | 168 |

### Procedure

The CoCoPIP survey comprises a mixed-methods approach, in which both quantitative and qualitative data was collected. This survey is logic-dependent and adaptive, only showing questions relevant to the parent’s current situation (i.e., first trimester/second trimester/infant aged 0-3/3-6 months). For the full survey, response time was ∼30 minutes and respondents were included in a £100 gift card prize draw. Here we focus in on qualitative survey data that addresses families’ experiences during pregnancy. As part of this survey parents or caregivers were asked to complete a structured assessment on access to healthcare (see *Table 2* for questions), alongside semi-structured questions focussed on their experience of healthcare support and access during their pregnancy (see *Table 3* for questions).

**Table 2:** Questions about healthcare access and appointment changes in response to the COVID-19 pandemic.

| Question | n (%) |
| --- | --- |
| **Have you attended some or all of your midwife, doctor or OB-GYN pregnancy appointments in person?** |  |
| Yes | 437 (86.5) |
| No | 68 (13.5) |
| Missing | 2 |
| **Do you feel comfortable attending your pregnancy appointments?** |  |
| Yes | 329 (65.4) |
| A little | 125 (24.9) |
| Unsure | 27 (5.4) |
| Not at all | 22 (4.4) |
| Missing | 4 |
| **Have you been offered online, phone or video call midwife appointments?** |  |
| Yes | 230 (45.5) |
| No | 276 (54.5) |
| Missing | 10 |
| **Do you feel that talking to your midwife online has allowed you to ask the questions you've wanted to and made you feel at ease?** |  |
| Yes | 63 (15.1) |
| A little | 84 (20.1) |
| Unsure | 70 (16.8) |
| Not at all | 200 (48.0) |
| Missing | 90 |
| **How well supported do you feel by your midwife, doctor (OB-GYN) and other prenatal healthcare professionals during this time?** |  |
| Extremely | 84 (17.0) |
| Very much | 140 (28.3) |
| Somewhat | 174 (35.2) |
| Not very | 59 (11.9) |
| I did not feel supported at all | 37 (7.5) |
| Missing | 13 |

**Table 3:** Qualitative questions asked participants regarding their healthcare support and appointment experiences.

| Number | Question | n |
| --- | --- | --- |
| 1 | Could you tell us about the support from your health-care provider during pregnancy? | 439 |
| 2 | Do you feel comfortable attending your pregnancy appointments? | 481 |
| 3 | Do you feel talking to your midwife online has allowed you to ask the questions you wanted to and made you feel at ease? | 381 |
| 4 | If not already covered in the prior questions, please provide further information on your experience of COVID-19. | 478 |
| 5 | In your own words please tell us about these classes (online) how often did you attend and how useful did you find them? | 230 |
| 6 | Prior to birth, were you certain whether partners and or family members could be present for the birth if you like, let us know how this communication or advice from the hospital made you feel? | 181 |

### Analysis

The qualitative data was imported from Qualtrics via Redcap (Harris et al., 2009) into Nvivo 12 (QSR International) software. A single researcher (KG) became familiar with the qualitative data, generated initial nodes and subsequently collated these nodes into broader themes. The study used a modified transcendental phenomenological (TP) approach (Moustakas, 1994) (i.e., the study is transcendental interpretivist and inductive; however, the main practical method of analysis used was thematic analysis (TA)). The trustworthiness of this data was assessed using the following criteria: credibility, transferability, dependability and confirmability (Guba & Lincoln, 1989). To address credibility, KG spent a period of weeks reading, reflecting and rereading the data; to address transferability a thick description of the data has been provided. Dependability involved the development of a clear audit trail in terms of the description of the methods used and a clear presentation of the findings using quotations to demonstrate themes. The coding trail was also double checked by EA. Finally, confirmability was addressed by ensuring a clear presentation of participant responses, and by providing a clear rational for each step involved in the methods and analysis; furthermore, an additional researcher (EA) has conducted a reliability analysis of 25% of the data to support the themes identified.

Further analysis of the data involved a sentiment analysis. This was conducted manually as a quality control check of the automatic coding of ATLAS.ti software revealed low reliability (<25% of autocodes were considered accurate by KG). Each response was read and categorised as ‘positive’, ‘negative’ or ‘neutral’ by a single researcher (KG) and a cross check of 10% of the sentiment labels were conducted by a second researcher (EA).

## Results

Of the 507 participants who responded to questions regarding their healthcare appointments and support during pregnancy, while few reported changes to their in-person appointments (13.5%), a third reported discomfort attending these appointments (34.7%), and half of participants said that they felt unsupported by their healthcare professionals during their pregnancy (54.26%) (see *table 2*).

The qualitative analysis identified a number of themes in relation to the healthcare provision that was provided: its role in increasing expectant parents’ anxiety; changes to the support that was provided including rushed care; lack of empathy; fewer appointments; the move to virtual provision; inability to have partners present; requirement to use PPE. The results of the sentiment analysis showed that of the total responses across all questions (n = 1320), 21.8% expressed positive sentiment, 28.1% neutral and 50.1% negative. This is consistent with the responses provided to the closed-ended questions (*see table 2*).

### Anxiety in relation to health care support

The most common theme was anxiety, which was reported across all six questions (see Table 3). Coding of the data for specific mentions of anxiety or related synonyms found Q6 to elicit the highest number of references to anxiety (25% of respondents).

Attendance at appointments was one notable reason given for feelings of anxiety:

> *‘I was very anxious about attending appointments and didn’t want to go’ (Q2)*

Participants also explicitly cited a lack of support as resulting in anxiety:

> *‘Felt that NHS decisions were not well communicated to expectant mums so there was a lot of anxiety over rumours about what to expect during labour’ (Q1)*

Whilst anxiety was often explicitly referred to, there were other instances of distress and lack of emotional/mental support that were described with one participant stating:

> *‘Constant changing advice from different departments left me in tears most days’ (Q6)*

And

> *‘Had medical support but no emotional/mental support during a time of great uncertainty’ (Q6)*

### Changes to healthcare support

A major independent theme identified, as well as a contributor to anxiety (as described above) was perceived lack of support. This reported lack of support was related to a range of issues including rushed and fewer appointments, perceived lack of sympathy, lack of face-to-face interactions, the result of online interactions, and being without a partner.

### Rushed and fewer appointments

There was reference to fewer and rushed appointments. One individual stated for example:

> *“…I feel like a lot of appointments have been more rushed (I’m assuming due to limiting exposure time due to COVID, etc) and I literally have no idea what to expect, or what a lot of things even mean/are. I feel as though a lot of my concerns about my pregnancy are also shrugged off by midwives etc (although I don’t think this is particularly COVID related)” (Q1)*

Similarly, another individual highlighted confusion and poor communication:

> *“Especially towards the end of the pregnancy I did not feel supported - midwives confused or missed appointments, no proper support in the midwife center lines (calls not answered etc), midwife appointments were always in a rush.” (Q3)*

In addition, lack of antenatal healthcare classes was also referenced, for example:

> *“…as a first-time mum, I feel like I haven’t been very well supported… I have no idea about anything that would usually be covered in antenatal classes.” (Q1)*

### Lack of sympathy

Another reported concern across all questions with regards to healthcare appointments during COVID-19 was the perceived lack of sympathy from healthcare staff. This included descriptions of insensitive treatment by staff to patients who had had previous losses during pregnancy, and failure to fully address mental health concerns. For example, respondents who experienced the loss of previous pregnancies stated:

> *‘My experience of the hospital where concerns were dismissed by a registrar and notes were lost so I would [sic] turn up for appointments where they weren’t expecting me. I felt bullied by the registrar who told me I am incapable of having babies.’ (Q1)*

And:

> *‘Generally good but different midwife each time meant different levels of support. None sensitive to previous losses!’ (Q1)*

And:

> *‘Felt despite having mental health illness no one wanted to know during pregnancy’ (Q5)*

And

> *‘…I feel like there has been no support offered or available. Especially not from a mental health perspective’ (Q1)*

### Restrictions on partners attending healthcare appointments

The lockdown restrictions meant that many healthcare sites did not allow partners to join them during routine visits and ultrasound procedures, and the data suggests that this increased feelings of isolation and loneliness. For example:

> *‘Visiting [sic] the hospital is a harrowing experience, and without a partner there for support makes it so much worse’ (Q2)*

And

> *‘Prior to this pregnancy I’ve [sic] had 2 miscarriages, I’ve [sic] suffered with depression, anxiety and panic attacks as a result. Every scan feels like they are going to tell me I’ve [sic] lost baby again and my anxiety goes through the roof. Haven’t [sic] felt comfortable or happy once on my own’. (Q2)*

And

> *‘Disappointed that partners have been excluded from all aspects of the routine appointments too, so I feel very alone on the journey.’ (Q2)*

Being alone, not only caused anxiety for participants, but in some cases, expectant mothers expressed reluctance to attend appointments:

> *“Reluctant to attend maternity triage or additional appointments as had to go alone” (Q2)*

### Lack of face-to-face meetings

A further impact of the changes to service delivery resulting from the COVID-19 pandemic was a decrease in the expected face-to-face contact with healthcare staff. Participants reported that interactions felt less personal and limited due to social distancing and specifically, the inability to see healthcare staff in person. One individual reported:

> *“I feel nervous about lack of face-to-face appointments. I have been having at home visits from an independent Midwife. Our first son was stillborn at 22 weeks, so I feel I need face to face appointments to check the baby and me.” (Q1)*

Some participants explicitly cited the lack of availability of face-to-face appointments as being a factor in their anxiety:

> *“A lot of my anxieties would be relieved by being able to see medical professionals face to face” (Q1)*

And

> *‘Lack of personal interaction has been detrimental to my pregnancy preparation and anxiety’ (Q1)*

Some respondents described not feeling able to ‘communicate fully’ or be properly examined using the phone or online classes:

> *‘It is hard to make contact on the telephone, you cannot communicate fully if you can’t use body language as well. Also, the midwife cannot see you so cannot examine you properly’ (Q3)*

And:

> *‘Due to my hearing disability, I find it incredibly difficult to communicate over online classes as I cannot always hear or lip read successfully.’ (Q5)*

The feeling of impersonality that resulted from the use of digital contact, was sufficiently anxiety inducing for at least one participant to refuse the offer of such help:

> *‘I refused as I felt anxious about the whole thing and felt it was inpersonal [sic]’ (Q5)*

Some respondents also felt that telephone appointments were rushed:

> *‘I feel telephone appointments have been rushed, it was not always explained clearly what the next appointment I was required to make was for or with and the midwives managed to pass responsibility onto the mother by allowing us to ask questions instead of going through everything thoroughly.’ (Q3)*

And

> *‘I had one video call and it was shocking. The midwife wasn’t able to listen to me or answer my questions. He appeared to just want to get off the phone.’ (Q3)*

### Use of PPE

While some respondents found the use of PPE reassuring (e.g., *‘Staff wore protective equipment so I felt safe’)*, it was also experienced by participants as being an additional source of anxiety and/or discomfort:

> *‘Wearing a mask whilst pregnant is very hot and [sic] get faint.’ (Q2)*

And

> *‘Visits were more rushed than expected and more stressful due to masks during hot weather’ (Q2)*

Participants were also aware of staff failure to adhere to the rules about PPE, with one reporting:

> *‘Mostly comfortable although one who took blood pressure did not wear mask’ (Q2)*

Scarcity in terms of the availability of PPE was also cited as being part of the difficulty of knowing what was safe:

> *‘I was in my ninth month at the height of Covid. Info was erratic, PPE was scarce. Every visit it was hard to know what was safe.’ (Q2)*

## Discussion

Our results reveal the impact that the COVID-19 pandemic and the significant changes that were made to the provision of prenatal healthcare across the UK has had on expectant parent’s experiences of maternity services throughout 2020. Quantitative results from the survey showed that a third of respondents reported discomfort in relation to attending appointments, and just over a half of participants said that they felt unsupported by their healthcare professionals during their pregnancy. Qualitative data from six open-ended questions suggest that not only did some women feel unsupported by the healthcare system during their pregnancy as a result of these changes, but also that the changes may have actually contributed to their feelings of anxiety. These findings are consistent with those reported in a large national survey of UK community-based practitioners including midwives and health visitors, who also reported that they felt that vulnerable families in particular were inadequately supported, as a result of the changes made to the health care provision following the first national lockdown (Barlow et al., 2020). Furthermore, these rapid and uncertain changes in the provision of healthcare services also contributed to practitioners’ own feelings of stress and anxiety.

Whilst there was some concern about the actual risk of being infected with COVID-19, the most substantial number of reported anxieties appeared to relate to participants’ concerns regarding whether their partners would be present during the birth (see Q6, *Table 3*). There was notable concern with respect to both attending appointments alone and potentially giving birth in a similar way. The impact of being alone in this way is something that has been found by other studies (Chivers et al., 2020; Karavadra et al., 2020) as well as being reported in the media. While concerns about this nationally led to a campaign to include birth partners at all appointments, which eventually resulted in a review of the public health guidelines in England and Wales (NHS, 2020), at the time of writing, the situation had not been revised in Scotland, and in some local authorities these strict measures have been reimposed in the national lockdown that was imposed in early 2021 in the UK.

Our data also suggest that another source of anxiety for prenatal participants was a perceived lack of support within the healthcare system itself. While some of these factors such as the use of virtual methods of service delivery including the telephone and video, have been identified by other studies (Chivers et al., 2020; Davenport et al., 2020; Karavadra et al., 2020) the current study also identified factors such as lack of sympathy and rushed appointments.

Antenatal anxiety is associated with a range of adverse perinatal outcomes (e.g., premature delivery; low birthweight) (Grigoriadis et al., 2018) in addition to a range of negative child outcomes that can persist into late adolescence including an increased risk of child behaviour problems (Stein et al., 2014). Furthermore, recent findings from surveys using standardised self-report measures, suggest that levels of pregnancy-specific anxiety have increased significantly during the Covid-19 pandemic (Ahlers-Schmidt et al., 2020; Davenport et al., 2020), as a result of a range of factors linked to imposed governmental guidelines (e.g., social distancing). The findings of the current study suggest that changes to the healthcare system that were instigated in response to the pandemic may have contributed to such feelings of stress/anxiety.

Lastly, similarly to Chivers et al’s (2020) sentiment analysis, which found 63% of responses to have negative sentiment vs 37% positive, we also found a marked weighting away from positive comments (21.8%). Whilst our analysis has a lower proportion of negative sentiment relative to Chivers et al, it was still prevalent with half of our sample reporting negative sentiment overall and a further 28.1% reporting neutrally. Indeed, perhaps due to the three-way division of positive/negative/neutral (rather than just negative/positive) our own study found an even lower proportion of positive sentiment relative to Chivers et al (2020) when asked about experience of healthcare during COVID-19.

### Limitations

As data were collected between July – December 2020 participants experiences reflect a period of fluctuating COVID-related government and healthcare restrictions, from the most severe national lockdown measures to a combination of severe to mild national/local restrictions. Furthermore, as a result of the fact that this study was conducted as a voluntary online survey, we cannot confirm all responses were by expectant parents or exclude bias in respondents with either positive or negative experience of their pregnancy. Whilst we advertised this study nationally, the majority of participants were White; therefore, the results cannot be generalised to a more ethnically diverse population. However, this study is part of an ongoing longitudinal study observing the impact of COVID-19 on pregnancy, infant development and parental mental health and we hope we increase the diversity of our sample longitudinally. Another limitation is that of the six questions posed not every participant gave a response to each one. Finally, from a qualitative perspective, due to the online survey nature of the project, it was not possible to probe and question further by means of interviews to further elucidate the links between perceived lack of support and anxiety.

#### Implications for practice and research

The Covid-19 pandemic has had a significant secondary impact on expectant women, and qualitative data suggests that going forward there is an urgent need for pregnancy-based services to better address the unique health care needs of each pregnant women, to better avoid the type of prenatal stressors that can have an enduring effect on their unborn child (see also Chivers et al., 2020; Iqbal et al., 2020). This should include having an opportunity to address expectant mother’s individual concerns, to be seen in person by healthcare providers with their partner as a matter of course, and to not feel rushed through their appointment. Further research is needed in terms of the ways in which the virtual delivery of pregnancy-based services can be optimised to meet the specific needs of women during these unprecedented times, and beyond.

### Conclusions

Changes to antenatal support and healthcare appointments in response to governmental guidance with regard to social distancing has had an adverse effect on the experiences of many pregnant women in the UK, with some evidence to suggest that it may have directly contributed to feelings of stress/anxiety. These findings point to an urgent need to better address the unique health care needs of each pregnant woman going forward.

## Data Availability

Data generated and analysed for this publication is not currently publicly available due to ethical and privacy restrictions.

## Acknowledgements

We are extremely grateful to all those families who gave their time to participate.

## Author statement

**Ezra Aydin:** Conceptualization, Methodology, Investigation, Data Curation, Validation, Writing - Original Draft. **Kevin A. Glasgow:** Investigation, Data Curation, Formal Analysis, Visualisation, Writing - Original Draft. **Staci M. Weiss:** Conceptualization, Methodology, Writing - Review & Editing. **Topun Austin:** Methodology, Supervision, Writing - Review & Editing. **Mark Johnson:** Supervision, Funding acquisition, Writing - Review & Editing. **Jane Barlow:** Supervision, Writing - Review & Editing. **Sarah Lloyd-Fox:** Conceptualization, Methodology, Supervision, Funding acquisition, Writing - Review & Editing.

